# PheMIME: An Interactive Web App and Knowledge Base for Phenome-Wide, Multi-Institutional Multimorbidity Analysis

**DOI:** 10.1101/2023.07.23.23293047

**Authors:** Siwei Zhang, Nick Strayer, Tess Vessels, Karmel Choi, Geoffrey W Wang, Yajing Li, Cosmin A Bejan, Ryan S Hsi, Alexander G. Bick, Digna R Velez Edwards, Michael R Savona, Elizabeth J Philips, Jill Pulley, Wesley H Self, Wilkins Consuelo Hopkins, Dan M Roden, Jordan W. Smoller, Douglas M Ruderfer, Yaomin Xu

**Affiliations:** Department of Biostatistics, Vanderbilt University, Nashville, TN, USA; Posit PBC, Boston, MA, USA; Division of Genetic Medicine, Department of Medicine, Vanderbilt University Medical Center, Nashville, TN, USA; Psychiatric & Neurodevelopmental Genetics Unit, Center for Genomic Medicine, Massachusetts General Hospital, Boston MA; Center for Precision Psychiatry, Department of Psychiatry, Massachusetts General Hospital, Boston MA; Department of Statistics, North Carolina State University; Department of Biomedical informatics, Vanderbilt University, Nashville, TN, USA; Department of Urology, Vanderbilt University Medical Center, Nashville, TN, USA; Division of Hematology and Oncology, Department of Medicine, Vanderbilt University Medical Center, Nashville, TN, USA; Department of Obstetrics and Gynecology, Vanderbilt University Medical Center, Nashville, TN, USA; Center for Drug Safety and Immunology, Department of Medicine, Vanderbilt University Medical Center, Nashville, TN, USA; Institute for Immunology and Infectious Diseases, Murdoch University, Murdoch, Western Australia, Australia; Vanderbilt Institute for Clinical and Translational Science, Vanderbilt University Medical Center, Nashville, TN, USA; Department of Pharmacology, Vanderbilt University Medical Center, Nashville, TN, USA; Stanley Center for Psychiatric Research, Broad Institute, Cambridge, MA; Department of Psychiatry and Behavioral Sciences, Vanderbilt University Medical Center, Nashville, TN, USA

**Keywords:** PheWAS, multimorbidity, interactive network analysis, UK Biobank, interoperability and reproducibility, electrical health records (EHR)

## Abstract

**Motivation:** Multimorbidity, characterized by the simultaneous occurrence of multiple diseases in an individual, is an increasing global health concern, posing substantial challenges to healthcare systems. Comprehensive understanding of disease-disease interactions and intrinsic mechanisms behind multimorbidity can offer opportunities for innovative prevention strategies, targeted interventions, and personalized treatments. Yet, there exist limited tools and datasets that characterize multimorbidity patterns across different populations. To bridge this gap, we used large-scale electronic health record (EHR) systems to develop the Phenome-wide Multi-Institutional Multimorbidity Explorer (PheMIME), which facilitates research in exploring and comparing multimorbidity patterns among multiple institutions, potentially leading to the discovery of novel and robust disease associations and patterns that are interoperable across different systems and organizations.

**Results:** PheMIME integrates summary statistics from phenome-wide analyses of disease multimorbidities. These are currently derived from three major institutions: Vanderbilt University Medical Center, Mass General Brigham, and the UK Biobank. PheMIME offers interactive exploration of multimorbidity through multi-faceted visualization. Incorporating an enhanced version of associationSubgraphs, PheMIME enables dynamic analysis and inference of disease clusters, promoting the discovery of multimorbidity patterns. Once a disease of interest is selected, the tool generates interactive visualizations and tables that users can delve into multimorbidities or multimorbidity networks within a single system or compare across multiple systems. The utility of PheMIME is demonstrated through a case study on schizophrenia.

**Availability and implementation:** The PheMIME knowledge base and web application are accessible at https://prod.tbilab.org/PheMIME/. A comprehensive tutorial, including a use-case example, is available at https://prod.tbilab.org/PheMIME_supplementary_materials/.

Furthermore, the source code for PheMIME can be freely downloaded from https://github.com/tbilab/PheMIME.

**Data availability statement:** The data underlying this article are available in the article and in its online web application or supplementary material.

## 1. Introduction

The global rise of multimorbidity, the simultaneous occurrence of several diseases within an individual, poses a significant challenge to public health and healthcare providers (Pearson-Stuttard *et al*., 2019; Skou *et al*., 2022; Morris *et al*., 2021; Hounkpatin *et al*., 2022). In-depth understanding of intricate multimorbidity can uncover the complex interactions between diseases that affect their course, severity, and the patient’s treatment response (Rodrigues *et al*., 2022; Pati *et al*., 2017; McQueenie *et al*., 2020; Chong *et al*., 2018). Moreover, it may reveal shared molecular mechanisms among various diseases (Fabbri *et al*., 2015; Koch, 2021; Dong *et al*., 2021), offering opportunities for innovative prevention strategies, targeted interventions, and personalized treatments for patients with multimorbidities (Smith *et al*., 2021; Nicholson *et al*., 2019; Jakovljević and Ostojić, 2013; Kuan *et al*., 2023).

Large-scale electronic health record (EHR) systems, linked to comprehensive health and clinical data, have significantly enhanced the statistical power to examine robust multimorbidity patterns that mirror real-world scenarios (van Driel *et al*., 2006; Calvin *et al*., 2022; Dong *et al*., 2021; Hanlon *et al*., 2022; Hassaine *et al*., 2020; McQueenie *et al*., 2020). Network analysis has been a powerful approach to decipher complex multimorbidity patterns (Strayer, Zhang, *et al*., 2023; Fotouhi *et al*., 2018; Aguado *et al*., 2020). Our recent work has demonstrated the interoperability of EHR-based multimorbidities and multimorbidity networks when compared across distinct EHR systems using standardized diagnostic codes (e.g., ICD9 or ICD10) (Vessels *et al*., 2023; Strayer, Vessels, *et al*., 2023).

Integrating multiple EHR systems can significantly improve the generalizability and robustness of multimorbidity characterization by identifying commonalities and differences across various populations. However, current standards for quantifying, representing, and analyzing multimorbidity patterns, especially for phenome-wide analyses leveraging large-scale EHRs, are still lacking (Ho *et al*., 2022; Pearson-Stuttard *et al*., 2019).

In this study, we introduce the Phenome-wide Multi-Institutional Multimorbidity Explorer (PheMIME), an interactive web application developed using the R programming language and the Shiny library (Chang *et al*., 2023). PheMIME enables researchers to interactively explore, compare, and discover multimorbidity patterns using a phenome-wide multimorbidity knowledge base compiled from three major EHR databases: Vanderbilt University Medical Center (VUMC), Mass General Brigham Hospital (MGB), and UK Biobank (UKB) (Strayer, Vessels, *et al*., 2023). With the selection of a specific disease phenotype of interest, users can directly compare multimorbidities between different institutions, conduct a network analysis based on individual or multi-institution combined multimorbidity network, and visualize dynamic clustering of significant multimorbidities by applying filters.

PheMIME provides a rich, explorable view of the multimorbidity patterns and relationships among diseases across multiple institutions. It can facilitate the validation of novel disease associations, the discovery of robust disease multimorbidity patterns, and potentially reveal new insights for future investigation. We demonstrate the utility of our tool in a case study focusing on schizophrenia.

## 2. Materials and Methods

### 2.1 Data Integration and Multimorbidity Knowledge Base

For our analysis, we accessed three EHR databases that included individual-level data for 250,000 random patients each from the Vanderbilt University Medical Center (VUMC) and Massachusetts General Brigham (MGB) EHR systems, as well as data from 431,105 subjects in the UK Biobank (UKB). For UKB, in-hospital records field-41270, which records the distinct diagnosis codes (ICD 10) a participant has had recorded across all the hospital inpatient records in either the primary or secondary position, was used for the analysis. The ICD 10 codes were then mapped to phecodes (Denny *et al*., 2010; Carroll *et al*., 2014) where a phecode was denoted as an occurrence for an individual if it appeared at least once in a UKB subject’s collapsed record across all hospital inpatient records, or if occurred two or more times in a patient’s record for either VUMC or MGB. Logistic regressions adjusting for patient age at last recorded visit, sex, race, and the number of unique phecodes present in patients’ records were run for each pair of two phecodes with two conditions of either phecode A or phecode B regarded as the outcome and the other one treated as the independent variable (Aguado *et al*., 2020; Strayer, Vessels, *et al*., 2023). The averaged test statistic from the two regression analyses is then used as an estimate for the multimorbidity strength between the phecode pair A and B. Multimorbidity strengths of all pairs are subsequently calculated and used to construct a phenome-wide multimorbidity network, which represents an undirected weighted network with disease as nodes and disease-disease connections as edges, weighted by the multimorbidity strengths. In addition, Pearson correlation of the common multimorbidity patterns between each pair of phecodes is used as a similarity score, which was also used to generate another undirected weighted network using the similarity scores as weights with disease as nodes and connections as edges(Strayer, Vessels, *et al*., 2023). We call this a multimorbidity similarity network. We finally consolidate all the summarized data from three institutions into a database. This serves as a knowledge base to fuel PheMIME for phenome-wide multimorbidity analysis. Our recent work has demonstrated the high reproducibility of phenome-wide multimorbidity patterns when comparing between the VUMC and MGB EHR systems (Strayer, Vessels, *et al*., 2023).

### 2.2 Design Scheme

PheMIME incorporates five primary modules: (1) “Disease Selection” module facilitates an interactive phecode table where users can readily search, filter, and select a disease phecode of interest. (2) “Multimorbidity Consistency Inspection” module enables users to assess the overall consistency of multimorbidity strength measurements from the knowledge base and compare them across multiple institutions. Additionally, this module incorporates features that underscore significant multimorbidity strengths linked to the selected phecode of interest, aids in assessing their distribution amidst all multimorbidity measurements, and enables comparison across institutions. (3) “Multimorbidity Network Visualization” module presents interactive visual representations of the multimorbidity networks constructed based on the multimorbidity strength measurements and allows users to examine and compare these networks. By integrating a dynamic network visualization and clustering methodology called associationSubgraphs (Strayer, Zhang, *et al*., 2023), this module permits exploration of the network’s subgraph structures and dynamic clustering for any multimorbidity network from a single institution or an amalgamation of multiple institutions. Moreover, this module enables users to apply filters and emphasize any significant multimorbidities and investigate their interconnections and enriched subgraphs. (4) “Reproducible Multimorbidities Exploration” module provides an interactive environment for examining a customizable subset of significant and/or reproducible multimorbidities across the institutions based on the multimorbidity strength measurements hosted in the knowledge base. This interface allows users to visualize the interconnections among chosen phecodes and the enriched multimorbidity subgraphs within the combined multimorbidity networks. Furthermore, this module accommodates pairwise comparisons between all institutions. (5) “Multimorbidity Similarities Exploration” module, much like the preceding one, uses multimorbidity similarity measurements as the strength measurement. It permits visualization of interconnected phecodes and the multimorbidity subgraphs enriched in the combined multimorbidity similarity networks and enables pairwise comparisons between all institutions.

## 3 Implementation example

Here, we outline a use-case scenario of PheMIME, using phenome-wide multimorbidity patterns of schizophrenia for exploration and network analysis. Starting with phecode 295.10 (schizophrenia), PheMIME generates an interactive Manhattan plot, enabling users to select phecodes based on their pairwise multimorbidity strength, also known as comorbidity strength (Figure 1A). A scatter plot for comparing between two systems is also generated, helping users to select robust multimorbidities that exhibit reproducible patterns across systems (Figure 1A). The interactive features of the Manhattan and Scatter plots enable the selection and highlighting of a consistent set of the same phecodes, based on the magnitude (Manhattan plot) and consistency (Scatter plot) of disease multimorbidities. For instance, the Manhattan plot might reveal phecodes highly co-occurring with schizophrenia, whereas the scatter plot could highlight phecodes that are either highly consistent across institutions or remain exclusive to individual institutions. The data table in Figure 1B shows comorbid phecodes of schizophrenia, its description, disease categories and corresponding multimorbidity strengths among three institutions. This table is interactive with the Manhattan and Scatter plots, allowing users to add or remove phecodes by clicking on the rows in the table. If a disease multimorbidity exhibits both a large magnitude and high consistency across different systems, it strongly indicates a robust disease multimorbidity across the systems.

**Figure 1:**
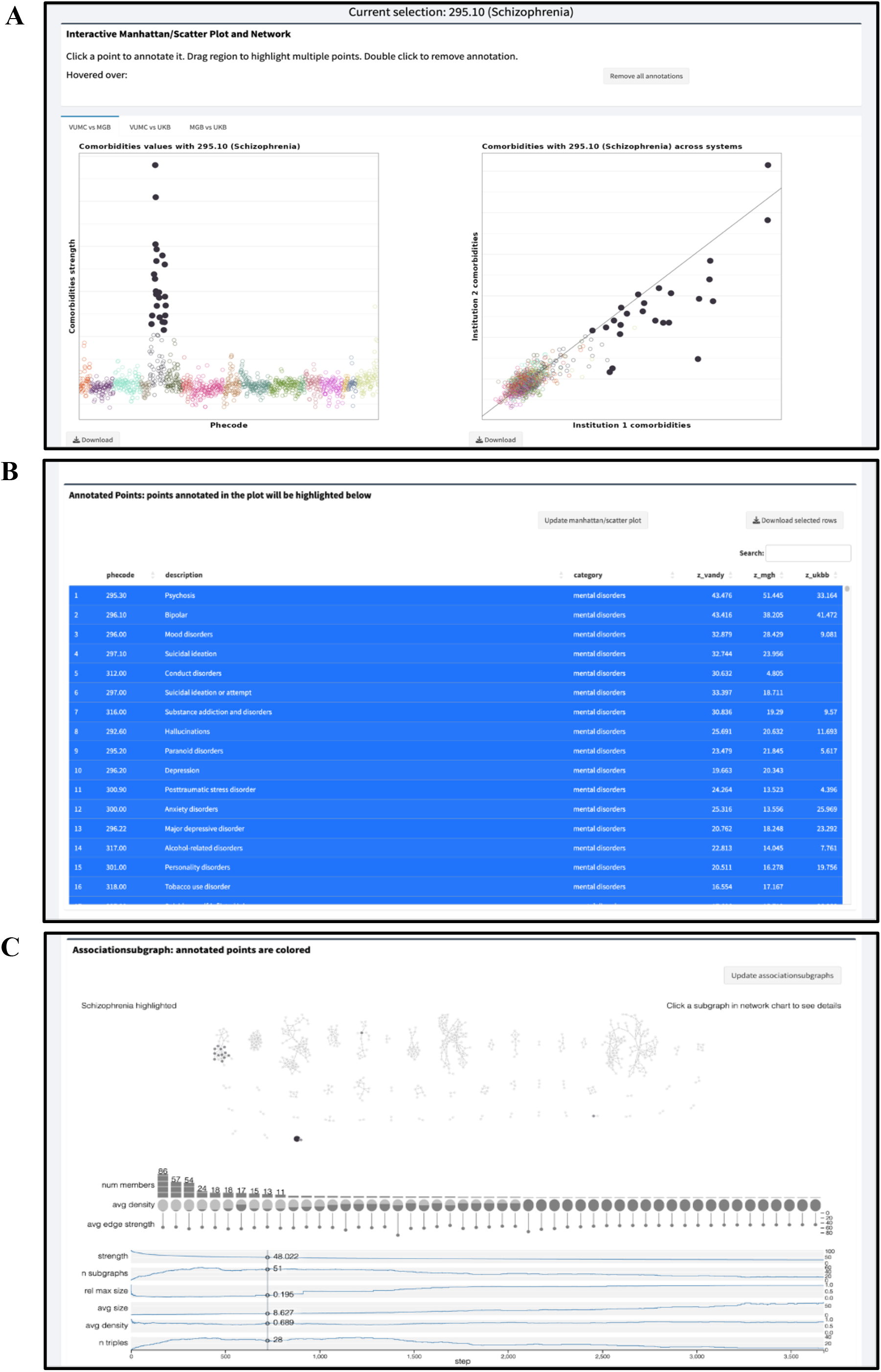
Comparison of disease multimorbidities in different populations. (A) Interactive Manhattan and Scatter plots comparing different cohorts, with schizophrenia (Phecode 295.10) selected as the phecode of interest. (B) Data table displaying comorbid phecodes associated with schizophrenia and the corresponding comorbidity strength values. The selected phecodes from part A are highlighted, and the table allows for user interaction with the plots. (C) Dynamic network analysis using associationSubgraphs, highlighting the user-selected phecodes from part A and their corresponding subgraphs.

For dynamic network analysis, the associationSubgraphs method (Strayer, Zhang, *et al*., 2023) has been enhanced to provide an interactive visualization to rapidly explore subgraph structures of schizophrenia multimorbidities in the selected phecodes that are both significant and reproducible across institutions (filled circles in Figure 1A and B). As shown in Figure 1C, network nodes are annotated into two groups, with the selected phecodes color-filled based on disease categories and the other unselected phecodes (nodes) color-filled in grey. As expected, a major subgraph enriching the selected schizophrenia multimorbidity phecodes is mental disorders, with other subgraphs showing enrichment in infectious diseases and neoplasms.

PheMIME also provides an expedient and accessible method for comparative analyses of multimorbidity patterns across different populations. Using schizophrenia again as an example, we discovered a strong comorbidity with viral hepatitis B and C (as detailed in the online materials), a finding in line with prior research(Leucht *et al*., 2007; Lluch and Miller, 2019). Yet, a detailed examination into multimorbidity patterns revealed more consistency of comorbidity within the patient cohorts of VUMC and MGB, with the comorbidity intensities being notably stronger in these cohorts as compared to the general UKB cohort, especially in the case of viral hepatitis B. This distinction could underscore heterogeneity due to population differences, including factors such as demographics and age restrictions in UKB. Given that the exact mechanisms behind viral hepatitis and schizophrenia comorbidity remain elusive, the observed disparities among different cohorts could motivate additional investigation regarding the mechanisms and impact of population differences on disease multimorbidity patterns.

## 4 CONCLUSIONS

We have introduced PheMIME, an interactive visualization tool specifically designed for analyzing multimorbidities across multiple EHR datasets, which simultaneously presents an extensive multimorbidity knowledge base consolidating data from three major EHR systems. Our intention is for users to utilize PheMIME to detect and extract meaningful disease multimorbidities, and to compare and validate them across a variety of institutions. To our understanding, PheMIME is the first knowledge base of its kind, integrating and comparing data from multiple extensive EHR systems while providing substantial support for efficient online analysis and interactive visualization to facilitate the discovery of complex multimorbidity patterns.

## Data Availability

All data produced in the present work are contained in the manuscript and in its online web application or supplementary material. Sharing of individual level data is restricted. The PheMIME knowledge base and web application are accessible at https://prod.tbilab.org/PheMIME/. A comprehensive tutorial, including a use-case example, is available at https://prod.tbilab.org/PheMIME_supplementary_materials/. Furthermore, the source code for PheMIME can be freely downloaded from https://github.com/tbilab/PheMIME.

https://prod.tbilab.org/PheMIME_supplementary_materials/

https://github.com/tbilab/PheMIME/

## Acknowledgement

We are deeply grateful for the insightful discussions with Kedir N. Turi, Sharon E. Phillips, Xiaopeng Sun, Lydia Yao, Eric Chen, Brian Sharber and Matt Krantz, which significantly enriched the development and direction of this work. Their input and expertise were invaluable and greatly appreciated.

## Author Declarations

All relevant ethical guidelines have been followed and any necessary IRB and/or ethics committee approvals have been obtained.

Yes

All necessary patient/participant consent has been obtained and the appropriate institutional forms have been archived.

Yes

Any clinical trials involved have been registered with an ICMJE-approved registry such as ClinicalTrials.gov and the trial ID is included in the manuscript.

Not Applicable

I have followed all appropriate research reporting guidelines and uploaded the relevant Equator, ICMJE or other checklist(s) as supplementary files, if applicable.

Yes

